# A CROSS-SECTIONAL STUDY OF THE PSYCHOLOGICAL IMPACT OF COVID-19 ON MEDICAL STUDENTS DURING & AFTER RECOVERY FROM COVID-19

**DOI:** 10.1101/2024.05.28.24308043

**Authors:** Anushka Misra, Damini Rane, P. Arun Bhat

## Abstract

**Introduction:** Mental health of medical students has been an utmost topic of discussion. It is important to know whether the upcoming doctors are mentally equipped to handle the pressure that comes with the profession. There is substantial literature indicating negative impact on the mental health during COVID-19 pandemic. Studies regarding the same on frontline workers seems scarce. Therefore, it is essential to assess the prevalence of symptoms anxiety and depression among medical students during and after the recovery from COVID-19 infection.

**Materials and methods:** It is a cross-sectional time bound study on 32 medical undergraduate students with active COVID-19 infection. They were screened using Self-reporting Questionnaire 20 (SRQ). Hamilton Anxiety Rating Scale (HAM-A) and Hamilton Depression Rating Scale (HAM-D) were administered on those who screened positive on SRQ-20 to assess the severity of anxiety and depressive symptoms. These were administered during and after COVID-19 infection on virtual platform.

**Results:** In our study, 59.37% population was screened positive for mental health disorder during COVID-19 infection. 89.5% of the positively screened respondents had mild anxiety symptoms and 10.5% had moderate anxiety symptoms. 47.4% of screened respondents had mild depressive symptoms, 31.6% reported moderate depressive symptoms and 21.1% had severe depressive symptoms. Following recovery, there was a reduction in severity of anxiety and depressive symptoms which was statistically significant (p<0.001).

**Conclusion:** Medical students have a propensity to develop anxiety and depressive symptoms while suffering from COVID-19 infection. Some may continue to have residual anxiety and depressive symptoms after recovery but the severity significantly reduces.

## BACKGROUND

Coronavirus disease (COVID-19) was identified as an atypical form of severe acute respiratory syndrome (SARS) in Wuhan, China in December 2019 caused by novel coronavirus-2 (SARS-CoV-2)^1,2^. The rapid spread of COVID-19 pandemic was a global health concern that can potentially impact the citizens of all nations due to its devastating consequences^3^.

Although ample clinical attention and research focus has been towards the treatment of the physical effects of the disease, the psychological fallout of the virus has been not been thoroughly studied on the affected individuals during and after recovery from the infection. As observed during previous viral outbreaks like SARS and MERS, viral infection and the isolation and quarantine swiftly culminate into anxiety, depression and sleep disturbances^4, 5^. Various psychiatric symptoms have been reported among SARS survivors, including post-traumatic stress disorder (PTSD), depression, panic disorder, and obsessive-compulsive disorder (OCD) up to 1 to 50 months of follow-up^6^. Previous prospective studies suggested that the psychological distress due to any illness was a predictor of future health and disease outcomes^7^. However, the current belief is that the psychological fallout of the COVID-19 pandemic may surpass the previous outbreaks mainly due to the widespread of misinformation by the social media which further perpetuates the sense of fear caused by the virus^8,9^.

The SARS-CoV-2 pandemic poses an additional risk for medical students because they serve as frontline workers. They are more likely to have stress and anxiety and hence are more prone to develop depression, anxiety and sleep disorders^10^. Studies on junior doctors and medical students revealed anxiety and depression amongst the frontline workers. Some highlighted a higher propensity among females than males whereas few studies did not show a gender vatiation^11,12,13^. Suicidal thoughts were prevalent in school going students during active COVID-19 infection and even after the recovery. Many of them reported of marked depression, anxiety, stress, loneliness, social anxiety, lack of self-control and many more negative perceptions during this time^14^. Furthermore, there is increased worry and fear of their health and their loved ones, disturbed sleep patterns, trouble concentrating, dwindling social interaction due to quarantine and social distancing which contributed to their other worries during the COVID-19 pandemic^15^.

These medical students are naturally concerned, not only because of their health but also about the impact of infectious disease on their peers and the potential impact on their studies. Targeted interventions provided after diagnosis of anxiety or depression can improve the mental health of these students and can potentially alleviate the psychological impact of the novel coronavirus. To date, there have been limited studies on the psychological impact of the SARS-CoV-2 outbreak on the mental well-being of medical students during and after recovery from the illness. This study aims to measure the levels of anxiety and depressive symptoms among medical students during and after the recovery from COVID-19 infection.

## MATERIALS AND METHODOLOGY

### Participants

This was a cross-sectional study done on 32 students who were diagnosed with COVID-19 infection. It was a time-bound study for 9 months conducted in a tertiary care teaching institute on all medical students from Phase 1 to 4. A sample size was not pre-determined and was established upon active participation during the course of the study. Clearance from Institutional Ethics Committee (IEC) was obtained. Students who completed the online interview were recruited in the study. Those students who did not respond to the questionnaire or responded incompletely were excluded from the study.

### Tools

A semi-structured questionnaire was devised to obtain socio-demographic and COVID-19 related details of the students. Self-reporting Questionnaire 20 (SRQ-20)^16^ is a self-administered scale which was provided to all students who had an active COVID-19 infection and also consented for the study. This is a screening tool for assessing anxiety and depressive symptoms which was circulated on an online platform. Hamilton Anxiety Rating Scale (HAM-A)^17^ and Hamilton Depression Rating Scale (HAM-D)^18^ were administered on those who screened positive on SRQ-20 to assess the severity of anxiety and depressive symptoms. These were administered during and 6 months after recovery from COVID-19 infection on virtual platform.

### Procedure

Medical students from Phase 1 to 4 who were diagnosed with COVID-19 taken up for the study. Verbal and written informed consent were taken before including them in the study. All the data was collected on an online video-calling platform. Socio-demographic and COVID-19 related details were obtained through the proforma. Self-administered SRQ-20 scale was provided to screen for anxiety and depressive symptoms at the time of active infection. Those who scored 8 or more on SRQ-20 were screened positive and were further assessed for severity of anxiety and depressive symptoms using HAM-A and HAM-D during and 6 months after recovery from COVID-19 infection. On the basis of the available information, inference was drawn regarding the difference in levels of anxiety and depressive symptoms during and after COVID-19 infection.

### Statistical analysis

Statistical analysis was performed using Statistical Package for the Social Sciences (SPSS) version 23.0. The collected data was entered on a Microsoft Excel Sheet. Data was expressed in descriptive statistics using frequency and percentage. The pre/post comparison of HAM-A and HAM-D was done using Wilcoxon signed-rank test. P-value <0.05 was considered statistically significant.

## RESULTS

The questionnaire was distributed via virtual platform among 600 medical students. A total of 32 students responded of whom 53.1% were females. More than half (65.6%) were less than 23 years of age. Out of the respondents, majority were from third and fourth year which was 31.3% in either year. Most of them did not have a family history of psychiatric illness and had never consulted a psychiatrist. 4 of 32 participants had taken treatment for anxiety or depression in the past. Only 1 participant had a past history of COVID-19 infection. Majority of the respondents did not require in-patient care during current COVID19 infection. The socio-demographic details have been summarized in **Table 1**.

**Table 1:**
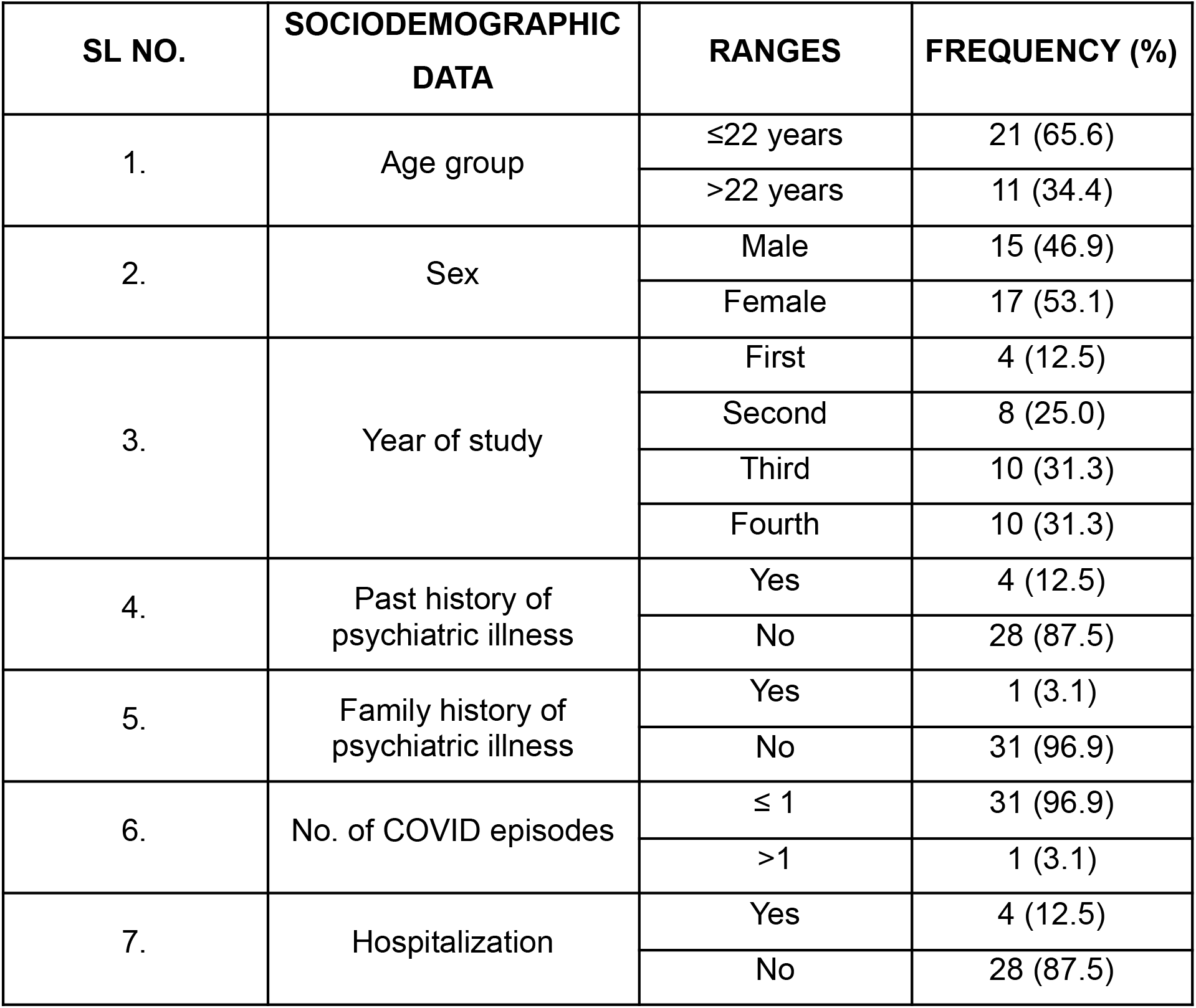
Sociodemographic data:

In our study, 59.4% were screened positive for mental health disorder with the mean age of 22 years (standard deviation [SD] =1.0) while 40.6% did not report of any mental health problems, mean age being 22.8 years (SD=1.67). Majority of the respondents were female (53.1%) from which 70.5% were screened positive for mental health disorder using the SRQ scale. Out of the remaining 46.9% male participants, less than half (46.6%) were screened positive for anxiety and depressive symptoms using SRQ scale. 15.8% of the participants with a positive SRQ score reported of a prior mental health disorder resulting in worsening of the symptoms. This data has been depicted in **Table 2**.

**Table 2:** Prevalence of mental health problems during COVID-19 infection:

| SL NO. | SRQ SCORES | FREQUENCY (%) |
| --- | --- | --- |
| 1. | <8 | 13 (40.6) |
| 2. | ≥8 | 19 (59.4) |

The subset of respondents that were screened positive for mental health related symptoms at the time of COVID-19 infection were assessed for severity of anxiety symptoms using HAM-A scale. 89.5% of the positively screened respondents had mild anxiety symptoms and 10.5% had moderate anxiety symptoms. Following recovery these respondents were reassessed to observe the change in severity of anxiety. Post recovery, 50% of these participants were found to have mild anxiety symptoms. This comparative data has been depicted in **Table 3** and **Figure 1**.

**Table 3:** Comparison of severity of anxiety symptoms during and after COVID-19 infection:

| SL NO. | HAM A SCORE INTERPRETATION | FREQUENCY (%) |  |
| --- | --- | --- | --- |
|  |  | DURING COVID INFECTION | POST RECOVERY |
| 1. | Mild | 17 (89.5) | 19 (100.0) |
| 2. | Moderate | 2 (10.5) | None |
| 3. | Severe | None | None |

**Figure 1:**
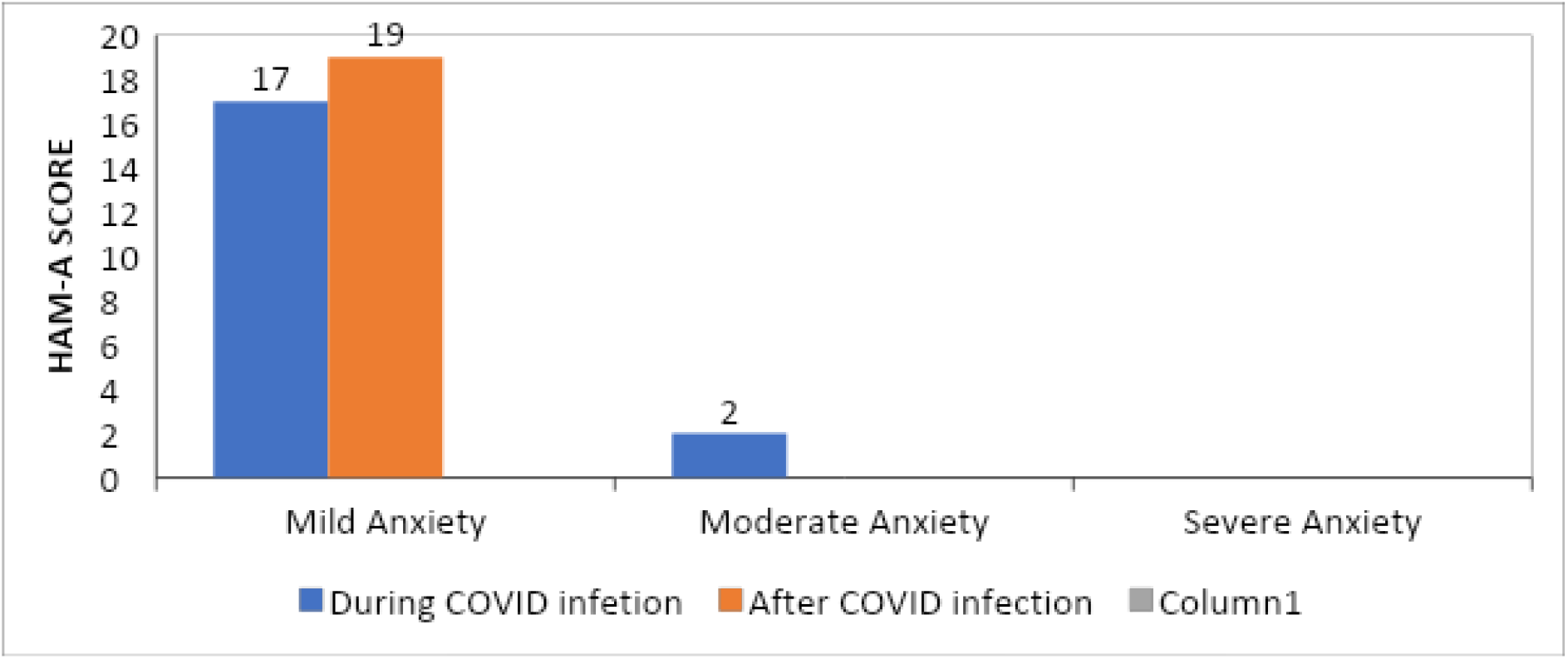
Comparison of severity of anxiety symptoms during and after COVID-19 infection:

Similarly, the respondents that were screened positive for mental health related symptoms were also assessed for severity of depressive symptoms using HAM-D scale. At the time of active infection, it was found that 47.4% of screened respondents had mild depressive symptoms, 31.6% reported moderate depressive symptoms and 21.1% had severe depressive symptoms. Additionally, they were reassessed for change in severity of depression after recovery. 10.5% participants reported to have mild depressive symptoms, 5.3% had moderate depressive symptoms and 5.3% had severe depressive symptoms respectively. This comparative data has been depicted in **Table 4** and **Figure 2**.

**Table 4:**
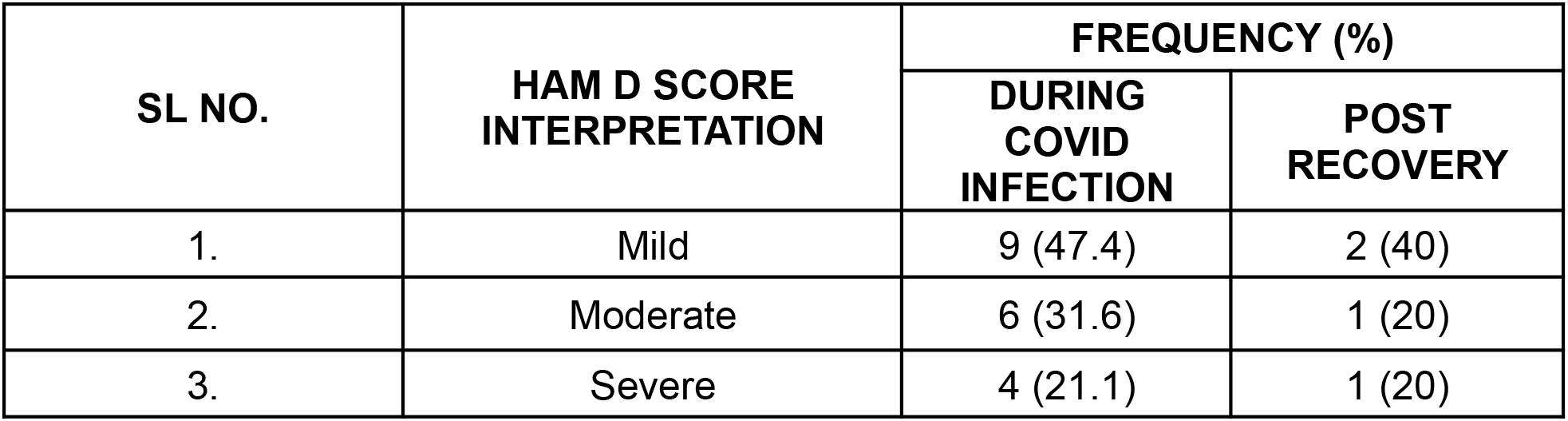
Comparison of severity of depressive symptoms during and after COVID-19 infection:

| SL NO. | HAM D SCORE<br>INTERPRETATION | FREQUENCY (%) |  |
| --- | --- | --- | --- |
|  |  | DURING<br>COVID<br>INFECTION | POST<br>RECOVERY |
| 1. | Mild | 9 (47.4) | 2 (40) |
| 2. | Moderate | 6 (31.6) | 1 (20) |
| 3. | Severe | 4 (21.1) | 1 (20) |

**Figure 2:**
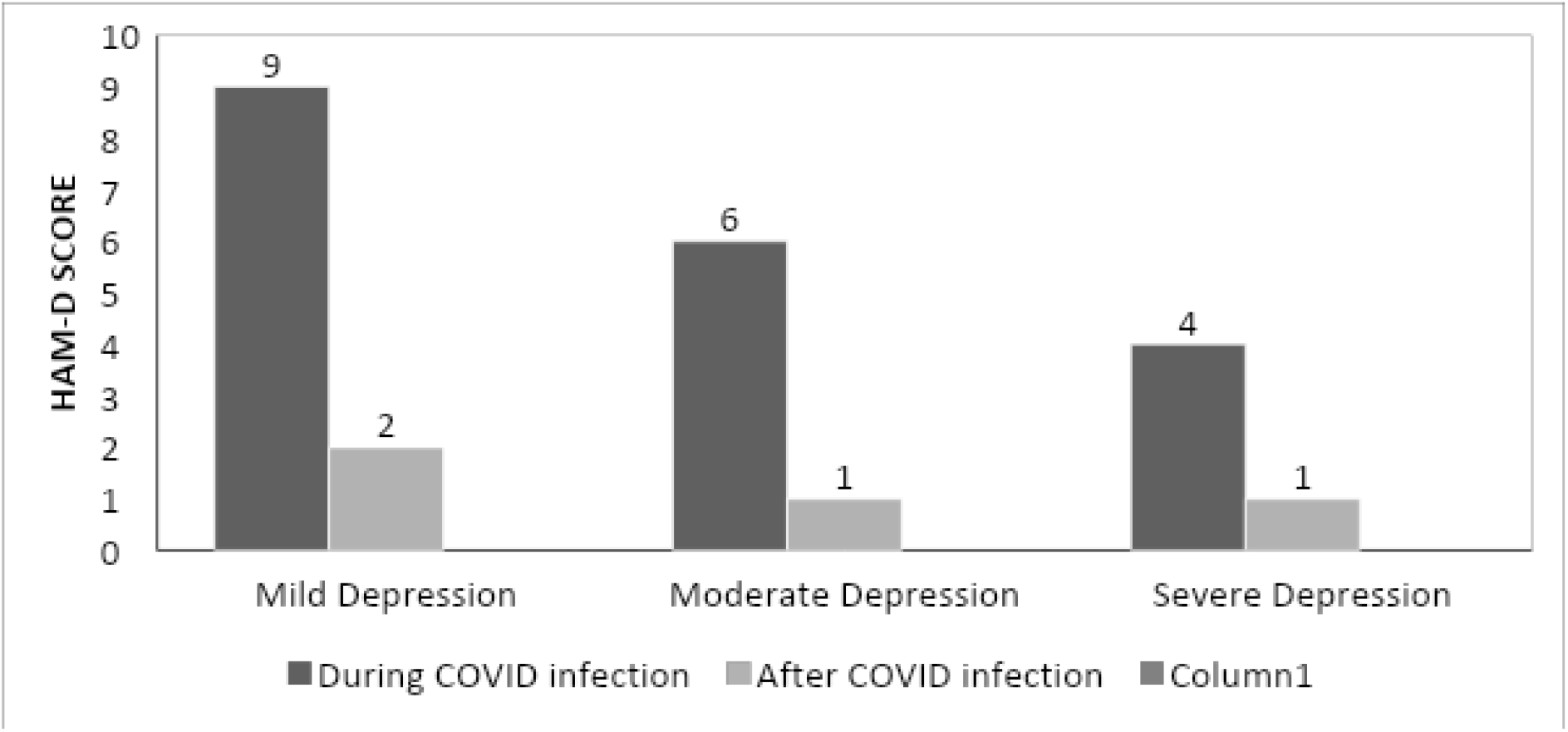
Comparison of severity of depressive symptoms during and after COVID-19 infection:

The anxiety and depressive symptoms that was precipitated during COVID-19 infection reduced after recovery from the infection and this reduction was found to be statistically significant (z=3.66, N=32, p<0.001). Anxiety symptoms reduced significantly with median HAM-A score during the infection being 5 [Interquartile range (IQR): 3-8] to post-infection score being 2 (IQR 1-8). Depressive symptoms also reduced significantly (z=3.83, N=32, p<0.001) with HAM-D score being 14 (IQR 11-17) during the infection and reducing to 9 (IQR 4-13) after recovery from infection.

## DISCUSSION

This cross-sectional study investigated mental health status of medical students during COVID-19 pandemic. The primary purpose was to investigate mental health status of medical students during and after recovery from COVID-19 infection and to determine their psychological wellbeing during the second wave of SARS-CoV2 virus.

Our study comprised of both genders but showed a higher number of women as compared to men. An Indian survey done in 2020-21 indicated that a higher percentage of women enrolled in medical science as compared to the men for higher education^19^. Furthermore, there has been an increase in female population in higher education from 2014 to 2020^19,20^. Hence our study population did not show a deviation from the general trend. The students in our study were within the age range of 20-26 years. Other studies showed a higher prevalence of anxiety and depression among younger population^21^. A survey looking into depression and anxiety among medical students indicated that both anxiety and depression were higher among third year medical students which was consistent with our study findings^22^.

There are studies done on Indian medical students which showed a predominance of anxiety and depression which was also seen in our study highlighting that 12.5% of the medical students had a previous history of psychiatric consultation^23^. Some of the participants in our study reported of anxiety and depressive symptoms during COVID-19 infection. Those who had a past history of mental health disorder reported of higher anxiety and depression during COVID-19 as compared to those who did not have a past history of psychiatric disorder. Studies contributing to our findings concluded that individuals with a history of mental health disorder have a higher prevalence of anxiety and depression^24^.

In our study, more than half the population with COVID-19 infection had anxiety and depressive symptoms which was found to be in line with the available literature suggesting that individuals suffering from COVID-19 infection experienced more adverse mental health outcomes than their non-COVID-19 counterparts^25,26^. Our study found an improvement in mental health status among undergraduate students after recovery from COVID-19 infection as compared to when they had COVID-19 infection. Our findings concerning the level of anxiety and depression symptoms among medical students were reported to be higher in females which complimented other studies which showed that females had significantly higher median anxiety and depression scores than the male participants^27^. With our study we noticed statistically significant decrease in anxiety and depression. Residual mental health consequences were noted even after recovery from the infection. This was in line with another study done on COVID-19 recovered patients from general population which highlighted that one-third of the population continued to experience anxiety and depression 6 months after the discharge^28^. Whereas, a similar study done on undergraduate students suggested no difference in mental health status before and after COVID-19 infection^29^.

## LIMITATIONS

The sample population for this study included undergraduate students pursing Bachelor of Medicine, Bachelor of Surgery. Therefore, the results obtained cannot be generalized to general population. Due to the nature of the study being a time-bound study, the sample size obtained was small making it further difficult to generalize the findings of our study.

## CONCLUSION

This study highlighted that undergraduate medical students have propensity to developing anxiety and depressive symptoms during COVID-19 infection. Certain socio-demographic groups were found of have been more at risk for mental health disorder; like female gender, respondents with past history of mental health disorder, students from third and fourth year of study. Moreover, respondents that had mental health disorder during COVID-19 infection had residual anxiety and depressive symptoms when reassessed after recovery. Nonetheless, there was a significant reduction in severity of their anxiety and depressive symptoms after recovering from COVID-19 infection.

## Data Availability

All data produced in the present study are available upon reasonable request to the authors

